# Short-term relaxation after cervical rotatory manipulation is more closely associated with somatosensory input than cracking sound: a randomized controlled EEG study

**DOI:** 10.64898/2026.06.13.26355570

**Authors:** Yuanxun Lin, Weixing Zhong, Meiling Yu, Yaoshuai yu, Lixin Tang, Fan Xue, Jundan Wei, Junhua Li, Yikai Li

**Author notes:** Corresponding author (JL); (YL).

## Abstract

**Background:** Cervical rotatory manipulation is commonly used for neck-related symptoms and is often accompanied by a cracking sound. This sound is frequently regarded as a sign of successful manipulation, but whether it contributes substantially to the immediate relaxation response remains unclear.

**Objective:** This study examined whether short-term relaxation after cervical rotatory manipulation is more closely related to manipulation-associated sensory input than to the cracking sound cue alone.

**Methods:** In this single-session, three-arm, parallel randomized controlled study, 54 healthy volunteers were allocated to cervical rotatory manipulation, sham manipulation, or sham manipulation plus simulated cracking sound. Subjective outcomes were assessed before and after intervention, including positive affect, negative affect, comfort, and satisfaction. Eyes-closed resting-state electroencephalography was recorded before and after intervention. Prespecified neural outcomes included frontal alpha power, frontal alpha/beta ratio, occipital individual alpha frequency, and alpha-band fronto-parietal and fronto-temporal functional connectivity.

**Results:** Cervical rotatory manipulation produced greater improvements in positive affect, comfort, and satisfaction than sham manipulation or sham manipulation plus simulated cracking sound, whereas negative affect remained generally stable across groups. These subjective responses were accompanied by short-term electroencephalography changes, particularly in frontal alpha/beta and alpha-band fronto-parietal and fronto-temporal functional connectivity. Changes in frontal alpha/beta ratio were positively associated with changes in positive affect. In contrast, simulated cracking sound alone did not reproduce the full subjective or electroencephalography response observed after real manipulation.

**Conclusions:** The immediate relaxation response after cervical rotatory manipulation appears to be more closely related to manipulation-associated sensory input than to the cracking sound cue alone. These findings provide preliminary neurophysiological evidence for distinguishing real manipulation effects from sound-related contextual cues.

**Trial registration:** The trial was registered with the International Traditional Medicine Clinical Trial Registry(URL: https://itmctr.ccebtcm.org.cn; ITMCTR2025001112).

## Introduction

Cervical rotatory manipulation (CRM) is a commonly used technique in traditional Chinese orthopedic manipulation for adjusting the functional state of the cervical facet joints^[1]^. Clinically, it is frequently applied for non-specific neck pain^[2]^, neck–shoulder discomfort^[3]^, dizziness^[4]^, and other cervical spondylosis-related symptoms^[5]^. From a biomechanical perspective, CRM shares characteristics with high-velocity, low-amplitude (HVLA) manipulation, in which a rapid, small-amplitude thrust is delivered after the targeted cervical segment reaches the elastic barrier, followed by immediate release^[6]^. In clinical practice, patients often report an immediate sense of “release” or relaxation after CRM, accompanied by reduced tension, increased comfort, and improved bodily experience. These immediate subjective changes are commonly considered an important component of the therapeutic effects of CRM^[7]^. However, the neurophysiological basis of the short-term relaxation response induced by CRM remains unclear. CRM is not a single stimulus but a complex sensory event involving therapist–patient contact, cervical segment positioning, cutaneous and soft-tissue somatosensory stimulation, cervical proprioceptive input, patient expectancy, and, in some cases, joint cavitation during the manipulation, which is commonly perceived as a cracking sound^[8]^. Patients and even some therapists often regard this sound as a sign of successful manipulation^[9]^. Nevertheless, whether the cracking sound itself makes a substantial contribution to the relaxation experience, or merely acts as a contextual cue that enhances patients’ expectations and treatment credibility, remains uncertain. Relying solely on subjective feedback after CRM may therefore confound genuine manipulation-related somatosensory modulation with sound-related cognitive or expectancy effects^[10]^. A rigorous experimental design is needed to distinguish the relative contributions of somatosensory input produced by CRM and the crack sound cue to the short-term relaxation response.

Resting-state electroencephalography (resting-state EEG) provides a non-invasive objective tool for evaluating short-term changes in relaxation, arousal, and emotional state^[11]^. In the present study, we focused on frontal alpha power, the frontal alpha/beta ratio, and occipital individual alpha frequency (IAF). Frontal alpha power is generally associated with lower cortical arousal, internally directed attention, sensory information inhibition, and relaxation^[12]^. During eyes-closed resting conditions, increased frontal alpha activity may serve as a potential neurophysiological marker of short-term relaxation or reduced tension. The alpha/beta ratio further reflects the relative balance between relaxation-related low-arousal activity and alertness-, tension-, or cortical activation-related activity^[13]^. Compared with alpha or beta power alone, the alpha/beta ratio may more intuitively characterize the transition from a relatively high-arousal state to a more relaxed state^[14]^. Occipital IAF reflects the peak frequency of the dominant individual alpha rhythm and is an important indicator of posterior dominant alpha activity during eyes-closed rest^[15]^. Unlike power-based measures, IAF primarily reflects the individualized frequency organization of alpha rhythms and resting-state brain functional characteristics, and may help determine whether CRM affects the baseline alpha rhythm state rather than merely altering local power intensity. In addition to spectral measures, we included alpha-band fronto-parietal and fronto-temporal weighted phase-lag index (wPLI) as functional connectivity measures^[16]^. wPLI can reduce, to some extent, the influence of volume conduction on phase synchrony estimation, and may help evaluate whether CRM modulates cross-regional functional coupling related to relaxation, attention, and emotional experience^[17]^. Therefore, frontal alpha power, the alpha/beta ratio, occipital IAF, and alpha-band wPLI connectivity can jointly characterize short-term neurophysiological changes after CRM from four complementary perspectives: local brain activity, arousal–relaxation balance, individualized alpha rhythm characteristics, and interregional coordination.

Previous studies on CRM or related cervical manipulation techniques have mainly focused on clinical analgesia, improvement in joint range of motion, or patient-reported outcomes. Although these outcomes are clinically important, they do not directly clarify whether a single manipulation session can induce measurable changes in relaxation-related brain activity. Moreover, few studies have experimentally separated the effects of real manipulation from those of the cracking sound. This distinction is particularly important because a simulated crack sound may increase the likelihood that participants believe they have received real CRM, but it cannot reproduce the cervical proprioceptive and deep somatosensory input generated by CRM. Therefore, determining whether CRM can induce subjective and EEG changes beyond the effect of the cracking sound may provide a more objective mechanistic basis for the immediate relaxation experience after CRM^[18]^.

To address this question, we conducted a single-session, three-arm, parallel randomized controlled EEG study in healthy volunteers. Participants were randomly assigned to the CRM group, sham group, or sham + simulated sound group. The three interventions were matched as closely as possible in body position, therapist contact, intervention duration, and standardized verbal instructions. In the sham + simulated sound group, a cracking sound was delivered through computer-controlled bone-conduction headphones to specifically test whether the cracking sound cue alone could reproduce the subjective and EEG effects of CRM. Eyes-closed resting-state EEG data and subjective ratings were collected before and after the intervention. We hypothesized that, compared with the sham and sham + simulated sound groups, CRM would produce greater improvements in positive affect, comfort, and satisfaction, accompanied by changes in relaxation-related EEG measures, particularly frontal alpha activity, the alpha/beta ratio, and alpha-band functional connectivity. In contrast, simulated cracking sound alone was expected to be insufficient to reproduce the full subjective experience and EEG response induced by CRM. By separating real CRM from the cracking sound cue, this study aimed to examine the relative contributions of manipulation-related somatosensory input and cracking sound cues to the short-term relaxation response following CRM.

## Materials and Methods

### Study design

This study was designed as a single-session, three-arm, parallel randomized controlled trial. Eligible participants were randomly allocated in a 1:1:1 ratio to one of three groups: CRM group, sham group, or sham + simulated sound group. Participants were blinded to the study hypothesis and outcome assessors were blinded to group allocation; however, full participant blinding to the presence or absence of an HVLA thrust was not feasible. The study was reported in accordance with the CONSORT statement, and the participant flow diagram is presented in Fig 1. The CONSORT checklist is provided in S1 Table.

**Fig 1.**
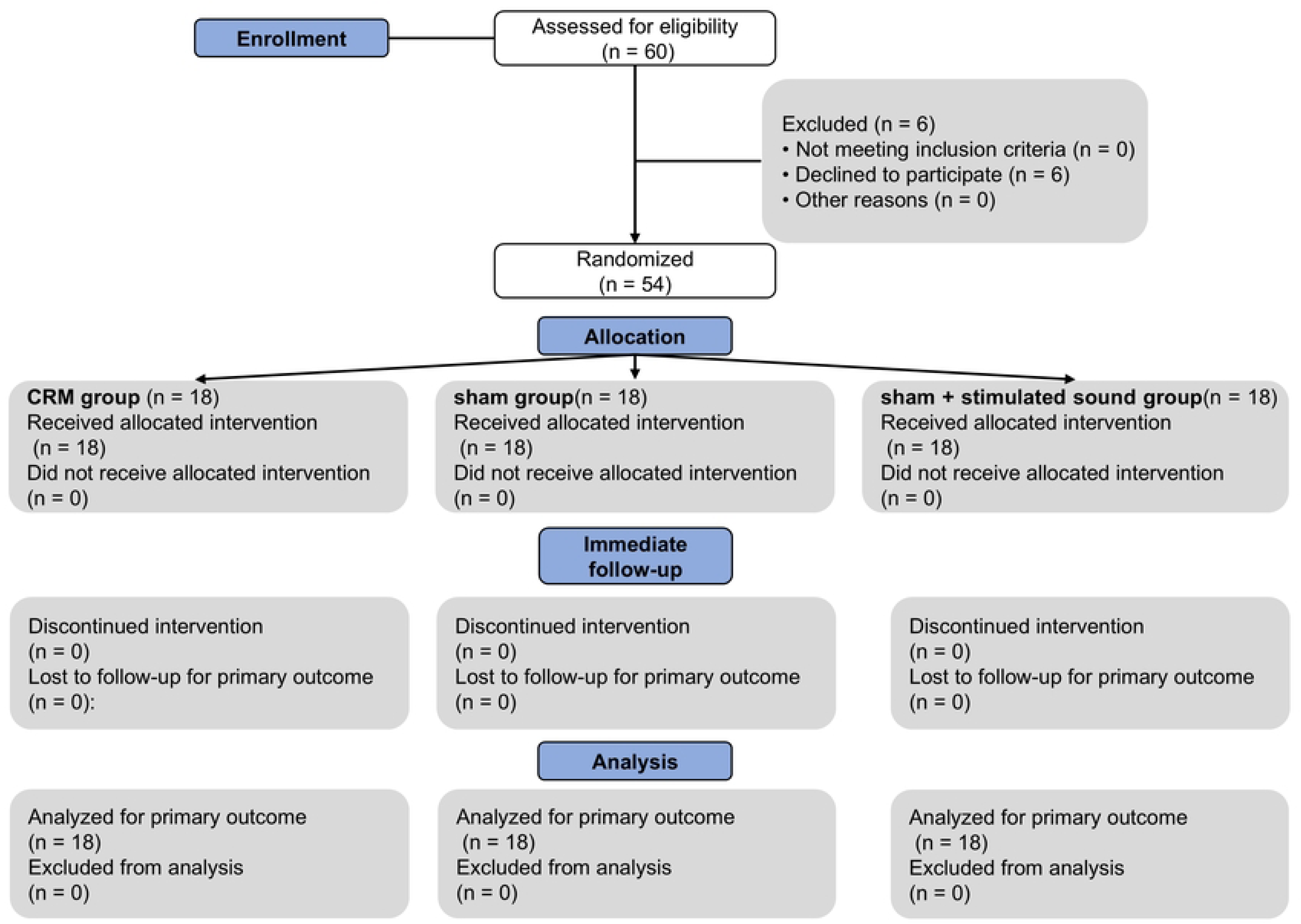
**CONSORT flow diagram of participant enrollment, allocation, follow-up, and analysis.**

### Participants

From April 2025 to August 2025, 60 healthy volunteers were recruited through advertisements posted in the Orthopedics and Traumatology outpatient department of The First Affiliated Hospital of GuangZhou Medical University. The study protocol was approved by the Ethics Committee of The Affiliated TCM Hospital of Guangzhou Medical University (Approval No.: 2025NK022; approval date: 7 March 2025). The trial was registered with the International Traditional Medicine Clinical Trial Registry (URL: https://itmctr.ccebtcm.org.cn Registration No.: ITMCTR2025001112; registration date: 23 March 2025). All participants provided written informed consent before enrollment.

The inclusion criteria were as follows: (1) age between 20 and 50 years; (2) no cervical disc herniation or other cervical structural lesions requiring treatment, as confirmed by imaging and clinical screening; (3) no history of tumor, major trauma, or cervical spine surgery; (4) body mass index (BMI) < 26 kg/m²; (5) Self-Rating Anxiety Scale (SAS) standard score < 53 and Self-Rating Depression Scale (SDS) standard score < 53; (6) no massage, manipulation, physical therapy, or related interventions within the previous month; and (7) willingness to participate and ability to provide written informed consent.

The exclusion criteria were as follows: (1) imaging evidence of cervical deformity or high-risk anatomical abnormalities, such as block vertebrae, os odontoideum, or basilar invagination; (2) severe cardiovascular or cerebrovascular disease, coagulation disorders, or other conditions that might increase the risk of cervical manipulation; (3) history of cranial or spinal trauma or surgery; (4) autoimmune or infectious diseases; (5) pregnancy or lactation; (6) implanted electronic devices, metal implants, or other conditions that might affect EEG acquisition or intervention safety; and (7) marked fear of manipulation or inability to cooperate with eyes-closed resting-state EEG recording.

Participants were allowed to withdraw from the study at any time by revoking informed consent. Investigators could discontinue a participant’s involvement if a serious adverse event occurred, if newly developed comorbidities or complications made continued participation inappropriate, or if the participant failed to complete EEG acquisition. Safety was monitored during the intervention and immediately after the intervention by proactive inquiry and direct observation. Recorded adverse events included, but were not limited to, neck pain or discomfort, dizziness, nausea, upper-limb numbness, transient neurological symptoms, and skin discomfort. For each adverse event, the onset time, duration, severity (mild, moderate, or severe), management, outcome, and relatedness to the intervention (definite, possible, or unrelated) were documented. In the event of a serious adverse event, the participant would immediately discontinue the intervention and receive appropriate medical assessment and management. The event would be reported to the ethics committee in accordance with institutional requirements, and the trial would be suspended or terminated if required by the ethics committee or the investigators’ safety assessment.

### Randomization, allocation concealment, and blinding

The randomization sequence was generated by an independent researcher using a computer-based random number generator with randomly permuted block sizes of 6 and 9 and a 1:1:1 allocation ratio. Allocation concealment was ensured using sequentially numbered, opaque, sealed envelopes prepared by a staff member who was not involved in participant recruitment, intervention delivery, outcome assessment, EEG preprocessing, or statistical analysis. After enrollment and completion of baseline assessments (P1), the therapist opened the next envelope in numerical order to determine the participant’s group assignment.

Outcome assessors and personnel responsible for EEG preprocessing and statistical analyses were blinded to group allocation. EEG datasets were coded before preprocessing and analysis. Because the HVLA thrust is perceptible, full participant blinding to the presence or absence of a thrust was not feasible. To minimize expectancy and sensory cues, the sham procedures were designed to match the CRM condition as closely as possible in participant positioning, therapist contact, contact points, procedural duration, and standardized verbal instructions, while omitting the HVLA thrust.

All participants wore the same Newman M110 bone-conduction headset (Shenzhen Haiguoda Technology Limited Company, Shenzhen, China) during the intervention procedure. In the CRM and sham groups, no sound was played through the headset. In the sham + simulated sound group, a cracking sound was delivered through the headset using a computer-controlled audio file. The playback file and intensity were fixed across participants based on pilot calibration to ensure audibility and comfort. At the end of the experiment, participants were asked whether they believed they had received CRM. Their responses were used for a descriptive assessment of blinding integrity. Due to the nature of the intervention, the therapist delivering the intervention could not be blinded; however, outcome assessors, EEG preprocessing personnel, and data analysts remained blinded to group allocation throughout data processing and analysis.

### Intervention

All interventions were delivered in a single session by a researcher with more than 5 years of clinical experience in cervical manipulation. Participants in all three groups wore the same Newman M110 bone-conduction headset during the intervention procedure. To balance the initial therapist contact across groups, all participants received an approximately 15-s manual contact procedure before the main intervention.

### CRM group

Participants were seated, and the operator stood behind them. Before the thrust procedure, a brief standardized soft-tissue release was applied for approximately 15 s. This release mainly consisted of acupoint-based pressing, kneading, and plucking techniques. The operator used the thumb to press and knead the paraspinal muscles around the cervical spinous processes. When tender points or palpable cord-like taut bands were encountered, the thumb was used to gently pluck the surrounding soft tissue. After the soft-tissue release, participants were instructed to flex the neck forward. The operator then placed the thumb of one hand against one side of the C5–C6 level, with the remaining fingers positioned on the contralateral side, while the other hand supported the participant’s mandible. The operator then applied slow upward traction and rotated the head approximately 45° to the left or right. After the restrictive barrier was reached, the operator briefly paused and delivered a controlled, rapid, small-amplitude thrust with an additional rotation of approximately 5°. This procedure could be accompanied by a cracking sound. No sound was played through the bone-conduction headset in the CRM group. No repeated thrust was performed solely to elicit a cracking sound.

### Sham group

The sham manipulation procedure was designed to match the CRM condition as closely as possible in participant positioning, therapist contact, contact points, procedural duration, and standardized verbal instructions. Participants were seated in the same position, and the operator stood behind them. The same bone-conduction headset was worn, but no sound was played. Before passive neck positioning, a matched 15-s light manual contact procedure was applied around the cervical paraspinal region. This procedure involved gentle contact only, without deep pressing, kneading, plucking of taut bands, or therapeutic soft-tissue release. The operator then placed the hands in positions similar to those used in the CRM group and gently positioned the participant’s neck within a comfortable range. However, the procedure did not approach the restrictive barrier, and no HVLA thrust was delivered. Only passive positioning and gentle stretching within a comfortable range were applied.

### Sham + simulated sound group

The sham manipulation procedure was identical to that used in the sham group, including the matched 15-s light manual contact procedure before passive neck positioning. During passive neck rotation, a brief standardized bone-conducted sound burst was delivered through the Newman M110 headset to mimic a cracking sound. Each burst lasted approximately 0.2–0.5 s and was delivered 1–2 times per session, synchronized with the peak rotation moment. The waveform and playback intensity were computer-controlled and kept constant across participants. The intensity was determined in a pilot calibration to ensure that the sound was clearly perceptible and comfortable. No HVLA thrust was delivered in this group.

### Time points and procedure

After a 15-min acclimatization period with eyes closed, baseline resting-state EEG was recorded for 3 min and defined as P1. To ensure procedural consistency across groups, the EEG device was removed after the pre-intervention recording in all participants and re-applied before the post-intervention recording. This procedure was necessary because the intervention involved contact with the occipital and cervical regions. The re-application process required approximately 3 min; therefore, the post-intervention EEG recording was operationally defined as an immediate post-intervention assessment but effectively began approximately 4 min after completion of the intervention. Post-intervention resting-state EEG was recorded for 3 min and defined as P2. Subjective outcomes included the International Positive and Negative Affect Schedule-Short Form (I-PANAS-SF)^[19]^, a modified version of the Massage Comfort Scale, and a satisfaction self-assessment form. The items, scoring methods, and interpretation of the comfort and satisfaction measures are provided in S3 Tables. Participants completed the I-PANAS-SF, the modified Massage Comfort Scale, and the satisfaction self-assessment form immediately before and after the intervention. To minimize the influence of environmental and procedural differences on EEG acquisition, the same recording workflow and standardized verbal instructions were used across all three groups. During EEG recording, participants were instructed to remain relaxed, keep their eyes closed, avoid unnecessary movement, and stay awake. Any instances of eye opening, swallowing, overt body movement, or drowsiness were documented during acquisition and used as references for artifact identification and rejection during preprocessing.

### Outcome measures and EEG processing

EEG was recorded using a 32-channel EMOTIV EPOC Flex system (Emotiv, California, USA). Electrodes were positioned according to the international 10–20 system, with bilateral ear electrodes used as references, yielding 30 EEG recording channels. Conductive gel GT5 (GreenTek, Wuhan, China) was used to reduce electrode impedance. All recordings were performed in a dedicated testing room under controlled environmental conditions. Electromagnetic interference, lighting, acoustic disturbance, and room temperature variation were minimized. Room temperature was maintained at 24 ± 2°C, lighting was kept dim, and humidity was maintained at 50 ± 5%. Participants were asked to wash their hair within 24 h before the experiment and were instructed to remain seated, relaxed, awake, and still during EEG acquisition. EEG data were preprocessed in MATLAB 2013b using the EEGLAB toolbox^[20]^. Detailed preprocessing parameters and procedures are provided in S4 and S5 Tables. Briefly, continuous eyes-closed resting-state EEG data were inspected and preprocessed to remove obvious artifacts, including eye opening, swallowing, overt body movement, and drowsiness-related segments. For each participant, EEG data at P1 and P2 were processed using the same pipeline.

The primary subjective outcome was the change in positive affect from P1 to P2, assessed using the I-PANAS-SF and expressed as a change score: Δ = P2 − P1. Secondary subjective outcomes included changes in negative affect, comfort, and satisfaction. Comfort and satisfaction were assessed at P1 and P2 using the modified Massage Comfort Scale and the satisfaction self-assessment form, respectively. EEG-derived outcomes included frontal alpha power, the frontal alpha/beta ratio, occipital IAF, and predefined alpha-band ROI-to-ROI wPLI connectivity metrics, including fronto-parietal (FP) and fronto-temporal (FT) connectivity.

Power spectral density (PSD, μV²/Hz) was estimated from preprocessed resting-state EEG data using Welch’s method. For each 3-min recording, 5-s buffers at the beginning and end were removed, and the remaining continuous data were segmented into non-overlapping 2-s epochs. PSD was estimated for each retained epoch using a Hamming window, and epoch-wise spectra were averaged to obtain channel-level PSD. The frontal region of interest (ROI) was defined a priori. PSD values were averaged across channels within the frontal ROI, and band power was calculated for the alpha (8–13 Hz) and beta (13–30 Hz) bands. The frontal alpha/beta ratio was calculated at each time point. Change scores were then computed for all spectral measures as Δ = P2 − P1. To characterize individual alpha rhythm features, IAF was estimated over the occipital ROI. Within the 8–13 Hz range, the frequency corresponding to the maximum PSD peak was identified as the IAF. IAF was computed separately for P1 and P2, and the change score was calculated as ΔIAF = P2 − P1. Functional connectivity was assessed using wPLI to reduce the influence of volume conduction on phase synchrony estimates^[21]^. Because the main hypothesis focused on relaxation-related alpha activity, inferential connectivity analyses were restricted to the alpha band. Preprocessed EEG data were filtered in the alpha band, and analytic signals were obtained using the Hilbert transform to extract instantaneous phase. wPLI was calculated for each channel pair within non-overlapping 2-s segments and then averaged across segments to generate individual whole-brain wPLI matrices at P1 and P2. To reduce the multiple-comparison burden and align with the a priori hypotheses, ROI-to-ROI connectivity metrics were predefined. Mean alpha-band wPLI was calculated for FP and FT connectivity by averaging wPLI values across all corresponding ROI channel pairs. Change scores for alpha-FP wPLI and alpha-FT wPLI were used for between-group comparisons and brain–behavior correlation analyses. Whole-matrix wPLI results were used for visualization and exploratory interpretation only.

### Sample size estimation

Based on our preliminary work, the sample size was estimated using G*Power 3.1 with a 1:1:1 allocation ratio, α = 0.05 (two-sided), and 90% power. The calculation indicated that at least 15 participants were required per group for the primary outcome, defined as the pre-to-post change in positive affect. To allow for possible nonparticipation before randomization and to improve the stability of exploratory EEG analyses, the target randomized sample size was increased to 54 participants, with 18 participants per group. In total, 60 volunteers were assessed for eligibility, six declined to participate before randomization, and 54 participants were randomized.

## Statistical analysis

All statistical analyses were performed using GraphPad Prism 9.0 and R 4.3.1 software. Continuous variables are presented as mean ± standard deviation (SD), and categorical variables are presented as counts and percentages. Baseline characteristics were compared among groups using one-way ANOVA, chi-square tests, or Fisher’s exact tests, as appropriate. For outcomes measured at P1 and P2, change scores were calculated as Δ = P2 − P1. Subjective outcomes were analyzed using linear mixed-effects models with group, time, and group × time interaction as fixed effects and participant ID as a random intercept. The group × time interaction was used to assess between-group differences in pre-to-post changes. For EEG-derived outcomes, group differences in Δ values were analyzed using one-way ANOVA or linear models, followed by pairwise comparisons when appropriate. Effect estimates were reported with 95% confidence intervals (CIs). Brain–behavior associations were examined using Spearman correlation analyses between changes in positive affect and changes in prespecified EEG outcomes. False discovery rate (FDR) correction using the Benjamini–Hochberg procedure was applied within related families of tests. Two-sided *P* values < 0.05 were considered statistically significant. An exploratory equivalence-oriented analysis was performed to examine whether the sham + simulated sound group differed meaningfully from the sham group. For each outcome, the between-group difference in change scores was calculated as Δsham + stimulated sound − Δsham, and the corresponding 90% CI was compared with prespecified smallest effect size of interest (SESOI) bounds using the two one-sided tests (TOST) procedure. Results were interpreted as equivalent when the 90% CI fell entirely within the SESOI bounds, not equivalent when it exceeded the bounds, and inconclusive when it overlapped the bounds. Bootstrap, permutation, and leave-one-out analyses were used as sensitivity analyses.

## Results

### Participant flow and baseline characteristics

A total of 60 healthy volunteers were assessed for eligibility. Six participants declined to participate, and 54 participants were randomized equally to the CRM group, sham group, or sham + simulated sound group, with 18 participants in each group. All randomized participants completed the intervention, post-intervention assessment, and primary outcome analysis. Baseline demographic characteristics were comparable among the three groups. No significant between-group differences were observed in age, weight, height, or BMI (all *P* > 0.05; Table 1).

**Table 1.** Baseline demographic characteristics of participants.

| Group | CRM<br>(n = 18) | sham<br>(n = 18) | sham + simulated sound<br>(n = 18) | *P*-value |
| --- | --- | --- | --- | --- |
| Age<br>(years old) | 24.17 ± 3.54 | 23.70 ± 3.40 | 23.81 ± 2.24 | 0.548 |
| Weight<br>(kg) | 60.16 ± 13.86 | 63.01 ± 15.71 | 62.11 ± 15.40 | 0.341 |
| **Height**<br>(m) | 1.67 ± 0.08 | 1.68 ± 0.08 | 1.67 ± 0.07 | 0.262 |
| **BMI**<br>(kg/m<sup>2</sup>) | 23.93 ± 3.93 | 23.49 ± 4.01 | 23.85 ± 3.76 | 0.584 |

Data are presented as mean ± SD. P values were obtained from one-way analysis of variance.
\*Significant effect ( $P < 0.05$ ).

### Blinding assessment

Participants’ beliefs regarding whether they had received CRM are shown in Table 2 and Fig 3A. In the CRM group, 14 participants believed that they had received CRM, 3 were unsure, and 1 denied receiving CRM. In the sham group, 10 participants believed that they had received CRM, 6 were unsure, and 2 denied receiving CRM. In the sham + simulated sound group, 11 participants believed that they had received CRM, 2 were unsure, and 5 denied receiving CRM. These descriptive findings suggest that the sham procedures retained a certain degree of treatment credibility across the control groups.

**Table 2.** Participants’ beliefs regarding whether they received CRM.

| Group | CRM<br>(n = 18) | sham<br>(n = 18) | sham + simulated sound<br>(n = 18) |
| --- | --- | --- | --- |
| **Belief received CRM** | 14 | 10 | 11 |
| **Unsure received CRM** | 3 | 6 | 2 |
| **Deny received CRM** | 1 | 2 | 5 |

Data are presented as counts. Participants were asked after the intervention whether they believed they had received CRM. CRM, cervical rotatory manipulation.

### Primary and secondary subjective outcomes

The primary outcome was the change in positive affect from P1 to P2. A significant group × time interaction was observed for positive affect. Positive affect increased markedly in the CRM group, whereas only a small increase was observed in the sham group and a slight decrease was observed in the sham + simulated sound group. Pairwise contrasts showed that the increase in positive affect was greater in the CRM group than in both the sham and sham + simulated sound groups. The sham + simulated sound group also differed from the sham group, indicating that simulated sound did not enhance positive affect relative to sham manipulation. For negative affect, no significant group × time interaction was observed. Negative affect remained generally stable across groups. For comfort, a significant group × time interaction was observed. Comfort increased in all groups, but the increase was greater in the CRM group than in the two control groups. Satisfaction also showed a significant group × time interaction, with the greatest increase observed in the CRM group. Detailed values, model results, and pairwise contrasts are presented in Table 3 and Fig 2B–E.

**Fig 2.**
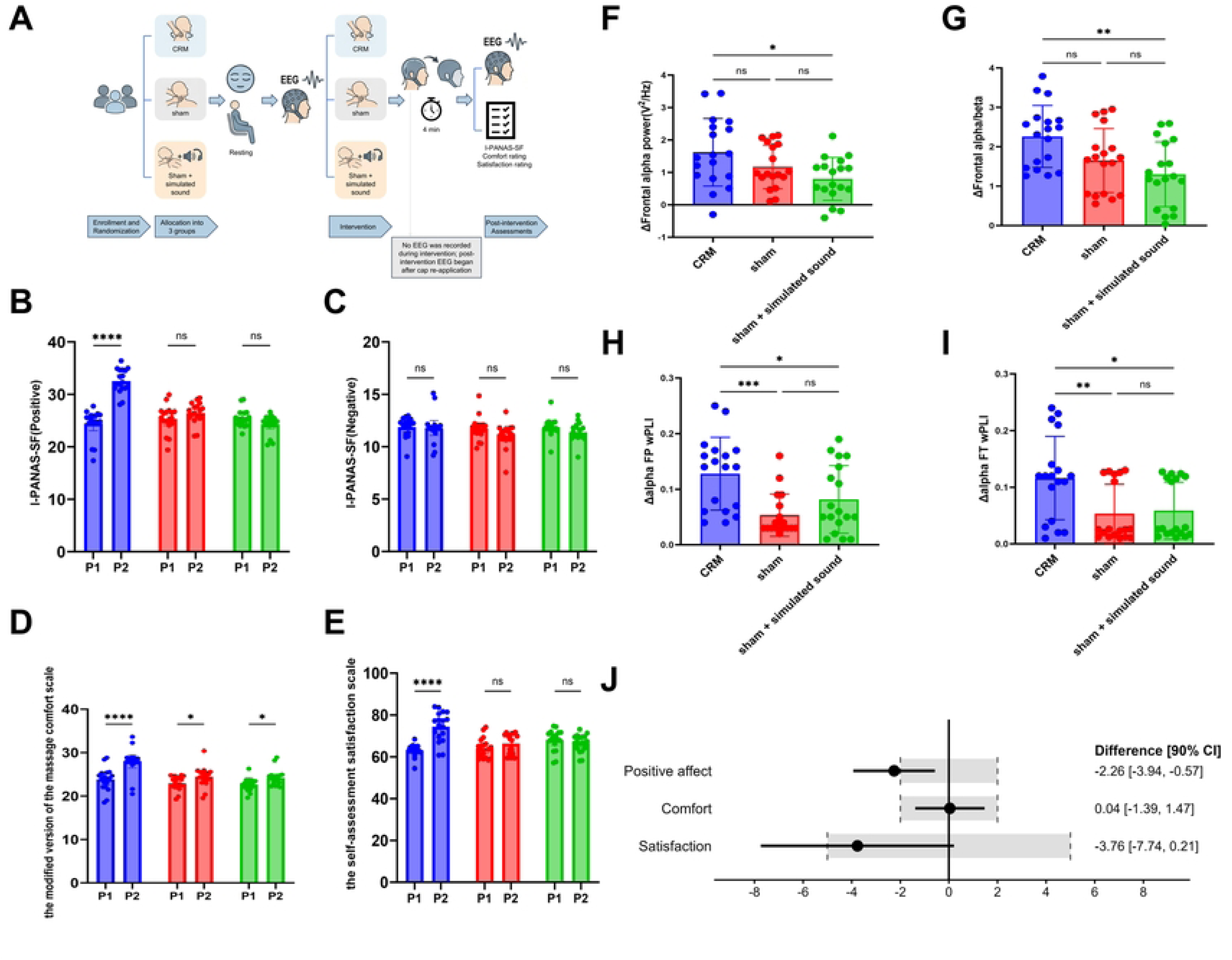
Study procedure and primary subjective and EEG outcomes. (A) Schematic illustration of the experimental procedure. Participants were randomized to CRM, sham manipulation, or sham manipulation plus simulated sound. Eyes-closed resting-state EEG was recorded before intervention (P1) and after intervention (P2). Because the EEG device was removed and re-applied for the intervention, the post-intervention EEG recording began approximately 4 min after the intervention. (B–E) Subjective outcomes at P1 and P2, including positive affect, negative affect, comfort, and satisfaction. (F–I) Changes in prespecified EEG-derived outcomes, including frontal alpha power, frontal alpha/beta ratio, alpha-band fronto-parietal wPLI, and alpha-band fronto-temporal wPLI. (J) Points and horizontal lines represent the estimated differences in change scores and their 90% confidence intervals for positive affect, comfort, and satisfaction between the sham + simulated sound group and the sham group. The vertical solid line at 0 indicates no between-group difference. The shaded regions indicate the prespecified SESOI bounds. Values to the left of 0 favor the sham group, whereas values to the right of 0 favor the sham + simulated sound group. CRM, cervical rotatory manipulation; EEG, electroencephalography; wPLI, weighted phase-lag index; FP, fronto-parietal; FT, fronto-temporal; CI, confidence interval. ns, not significant; \**P* < 0.05; \*\**P* < 0.01; \*\*\**P* < 0.001; \*\*\*\**P* < 0.0001.

**Fig 3.**
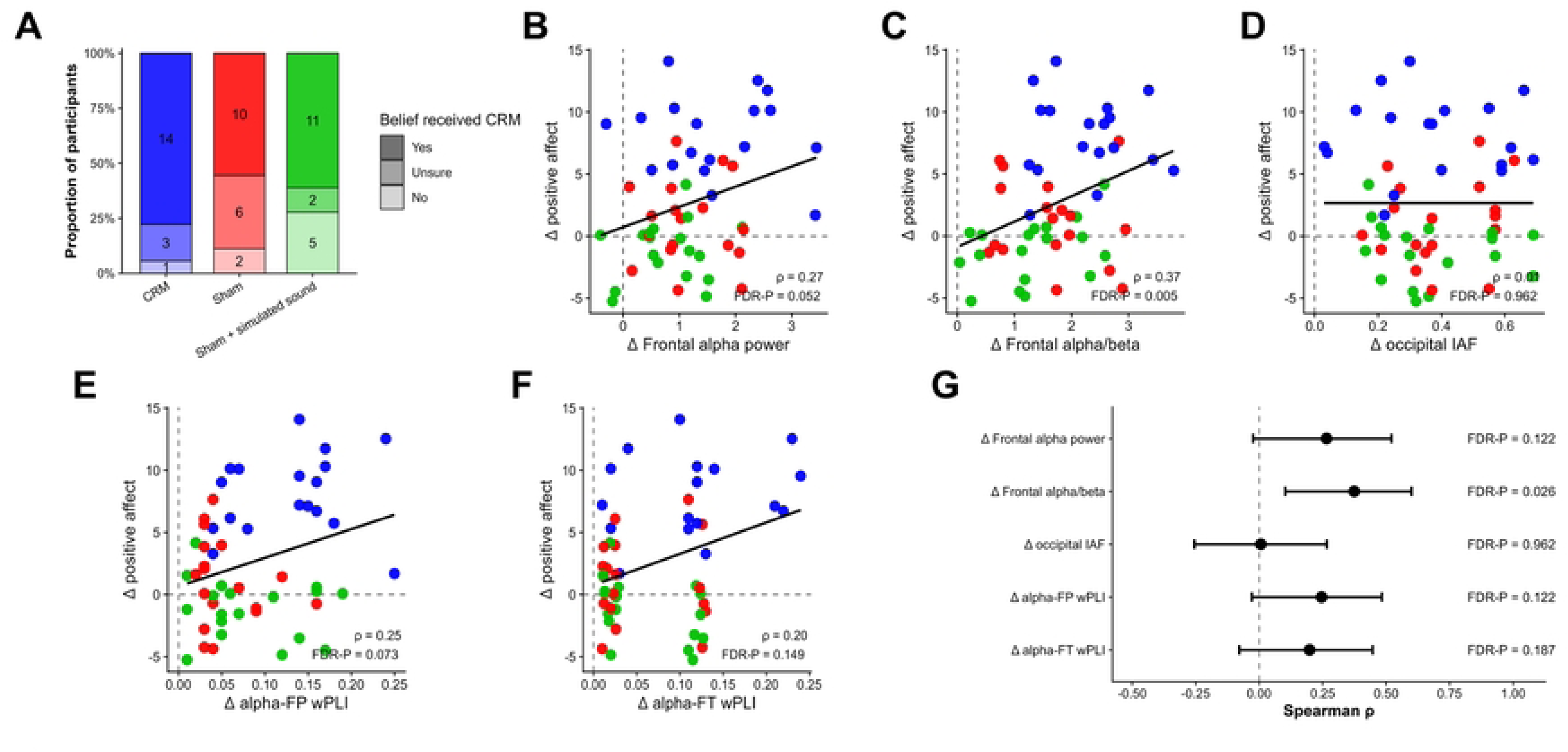
Blinding assessment and brain–behavior coupling analyses. (A) Participants’ beliefs regarding whether they received CRM after the intervention. (B–F) Scatter plots showing associations between changes in positive affect and changes in prespecified EEG-derived outcomes: frontal alpha power, frontal alpha/beta ratio, occipital IAF, alpha-band fronto-parietal wPLI, and alpha-band fronto-temporal wPLI. Each dot represents one participant, and colors indicate intervention groups. The black line represents a fitted linear trend for visualization only; statistical inference was based on Spearman correlation analysis. (G) Summary forest plot of Spearman correlation coefficients with corresponding FDR-adjusted P values. CRM, cervical rotatory manipulation; IAF, individual alpha frequency; wPLI, weighted phase-lag index; FP, fronto-parietal; FT, fronto-temporal; FDR, false discovery rate.

**Table 3.**
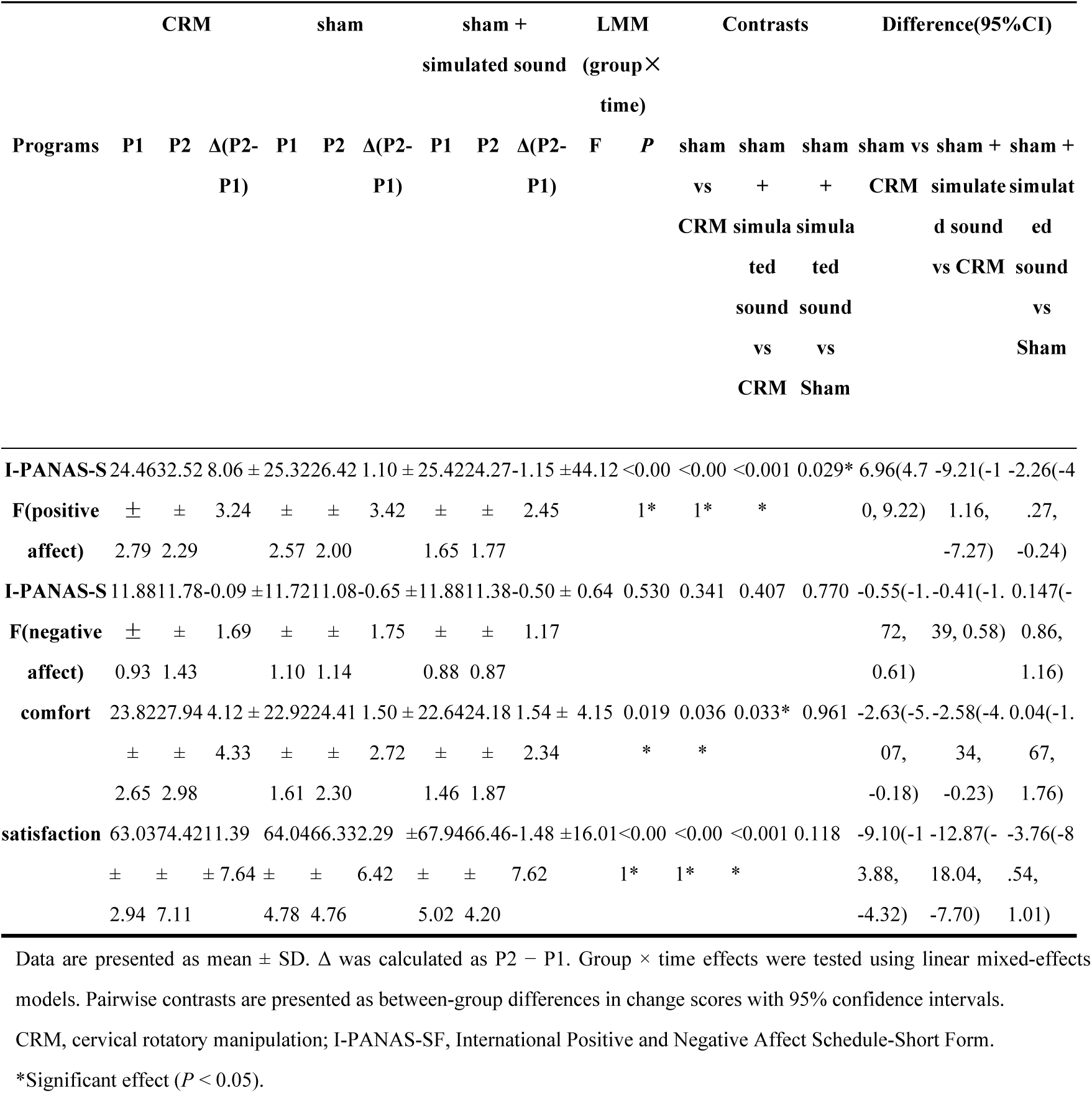
Primary and secondary subjective outcomes before and after intervention.

### EEG-derived outcomes

Changes in prespecified EEG-derived outcomes are shown in Table 4 and Fig 2F–I. The CRM group showed greater increases in frontal alpha power and frontal alpha/beta ratio than the sham + simulated sound group. The frontal alpha/beta ratio also differed between the CRM and sham groups. In contrast, occipital IAF did not differ significantly among the three groups. For alpha-band functional connectivity, significant group differences were observed in both fronto-parietal and fronto-temporal wPLI. The CRM group showed greater alpha-FP and alpha-FT wPLI changes than the sham group and the sham + simulated sound group. No significant differences were observed between the sham and sham + simulated sound groups for these connectivity outcomes. Whole scalp-level visualizations was used for exploratory interpretation and are shown in Fig 4.

**Fig 4.**
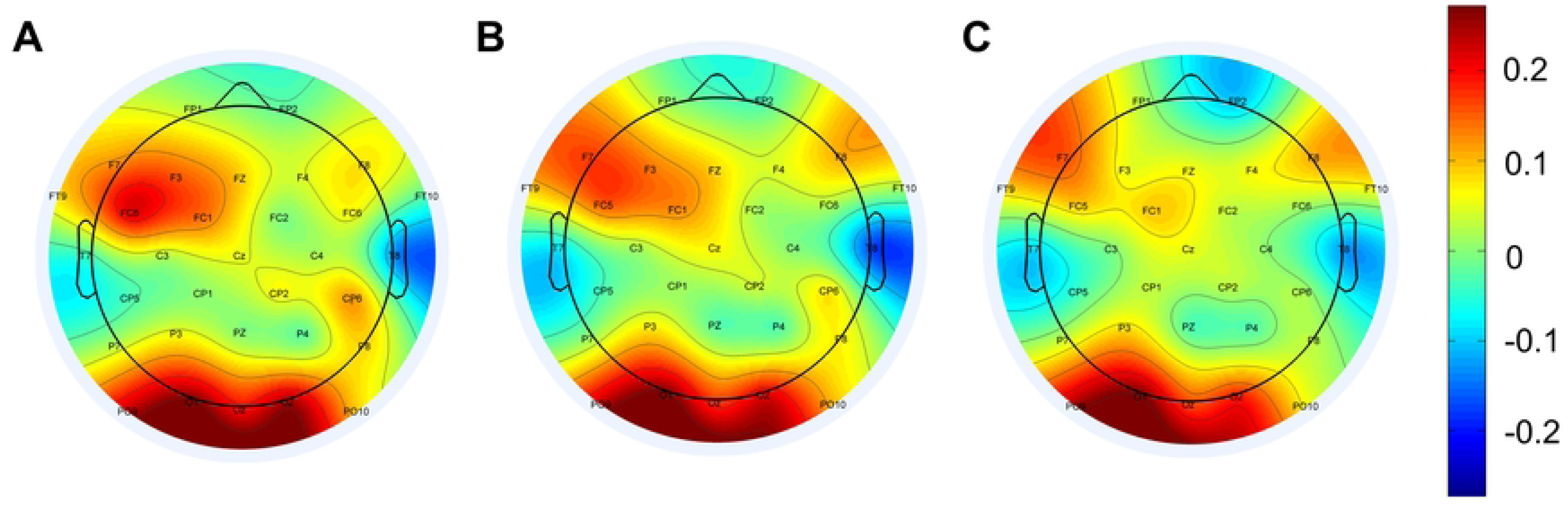
Scalp topography of alpha-band power changes after intervention. Scalp topographic maps show group-level alpha-band power changes from P1 to P2 (Δ = P2 − P1). (A) CRM group. (B) sham group. (C) sham + simulated sound group. Warm colors indicate relative increases in alpha power, whereas cool colors indicate relative decreases. The same color scale was applied across the three groups to facilitate visual comparison. The frontal region was used as the prespecified region of interest for statistical analysis. CRM, cervical rotatory manipulation.

**Table 4.**
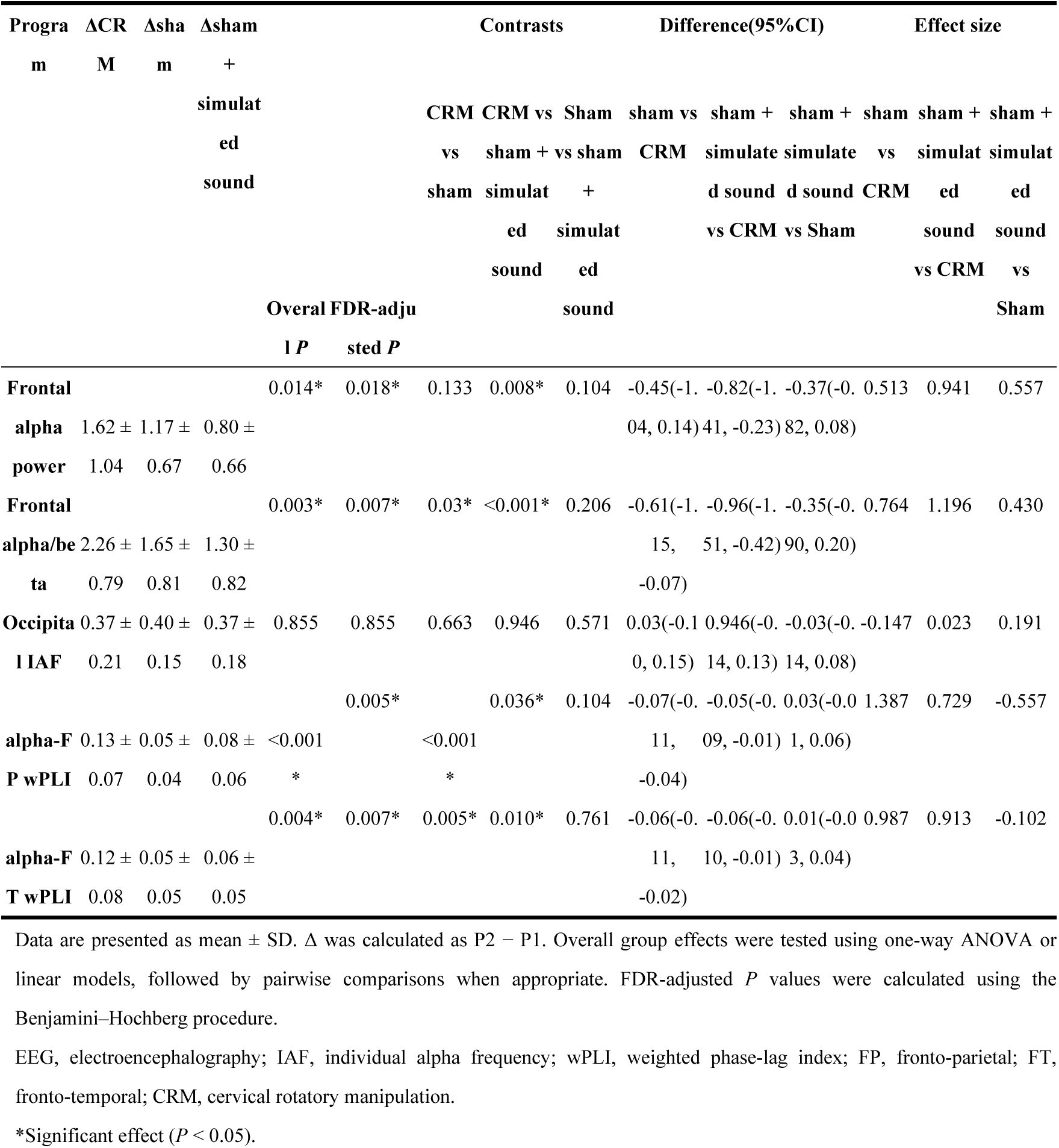
Changes in prespecified EEG-derived outcomes.

### Brain–behavior coupling

Spearman correlation analyses were performed to examine whether changes in positive affect were associated with changes in prespecified EEG outcomes. Changes in frontal alpha/beta ratio were positively correlated with changes in positive affect after FDR correction. Associations between positive affect and frontal alpha power, alpha-FP wPLI, and alpha-FT wPLI showed positive trends but did not survive FDR correction. No meaningful association was observed between changes in positive affect and occipital IAF. These results are shown in Fig 3B–G.

### Exploratory equivalence-oriented analysis of simulated sound

To examine whether simulated cracking sound produced a practically meaningful effect beyond sham manipulation, exploratory equivalence-oriented analyses compared the sham + simulated sound group with the sham group. For positive affect, the 90% confidence interval exceeded the prespecified SESOI bounds, indicating that the difference was not equivalent. For comfort and satisfaction, the results were inconclusive. For EEG-derived outcomes, most comparisons between sham + simulated sound and sham manipulation were inconclusive, with no clear evidence that simulated sound reproduced the EEG changes observed after CRM. Robustness analyses using bootstrap, permutation, and leave-one-out procedures are summarized in Table 5.

**Table 5.**
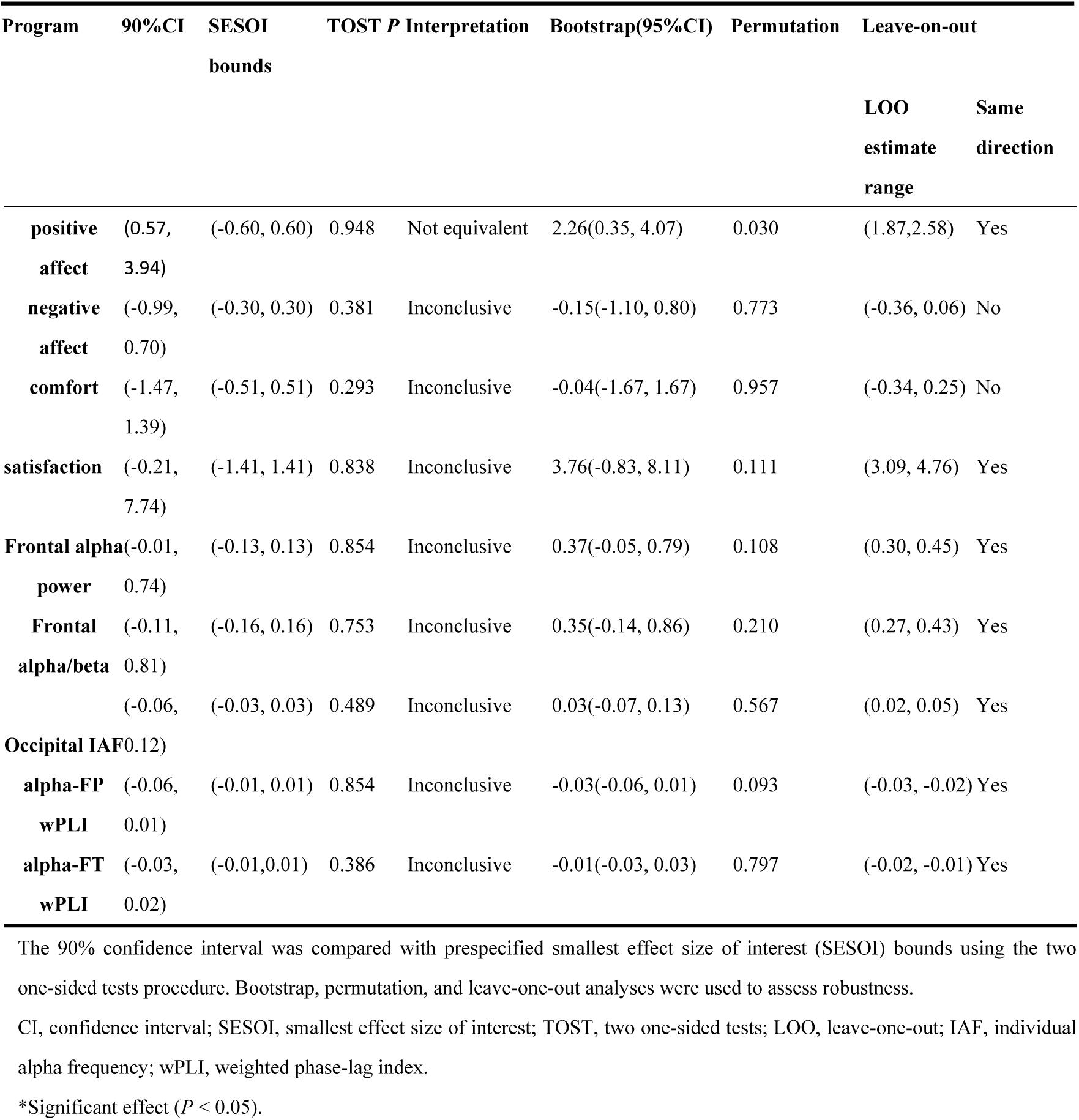
Exploratory equivalence-oriented analysis comparing sham + simulated sound with sham manipulation.

Overall, CRM induced stronger improvements in positive affect, comfort, and satisfaction than sham manipulation or sham manipulation plus simulated sound. These subjective effects were accompanied by changes in relaxation-related EEG markers, particularly frontal alpha/beta balance and alpha-band fronto-parietal and fronto-temporal connectivity. These findings support the interpretation that the short-term relaxation response after CRM is more closely associated with manipulation-related somatosensory input than with the cracking sound alone.

## Discussion

### Principal findings and interpretation

This single-session, three-arm randomized controlled EEG study examined whether the short-term relaxation response after CRM was more closely related to the manipulation procedure itself or to the cracking sound cue alone^[22]^. The main findings were that CRM produced greater improvements in positive affect, comfort, and satisfaction than sham manipulation or sham manipulation plus simulated cracking sound, whereas negative affect remained generally stable across groups. These subjective changes were accompanied by relaxation-related EEG modulation, particularly in frontal alpha/beta ratio and alpha-band fronto-parietal and fronto-temporal functional connectivity. In contrast, the sham + simulated sound condition did not reproduce the full subjective or EEG response observed after CRM.

These findings suggest that the immediate relaxation experience after CRM is unlikely to be explained solely by the auditory cracking sound^[23]^. In clinical practice, cracking sound is often regarded by patients and some therapists as a sign of successful manipulation^[9]^. However, in the present study, adding a simulated cracking sound did not produce CRM-like improvements in positive affect, comfort, satisfaction, or alpha-band EEG connectivity^[24]^. This result suggests that the simulated cracking sound alone was insufficient to account for the complete short-term response induced by real CRM. It should also be noted that the sham interventions produced modest changes in some subjective measures, especially comfort. This is reasonable because the sham procedures included therapist contact, cervical positioning, light manual contact, and gentle passive movement. These components may have nonspecific sensory and contextual effects. Nevertheless, the stronger response in the CRM group suggests that the HVLA thrust and the associated cervical sensory input provided additional effects beyond contact, positioning, and expectation alone^[6]^.

### EEG evidence and brain-behavior coupling

The EEG findings provide physiological evidence consistent with the subjective outcomes^[25]^. Among the spectral measures, frontal alpha power showed a group effect, whereas the frontal alpha/beta ratio demonstrated a clearer CRM-related pattern. Because increased alpha activity during eyes-closed rest has often been associated with lower arousal and relaxed internal attention, whereas beta activity is more closely related to cortical activation or alertness, the alpha/beta ratio may better reflect the balance between relaxation-related and arousal-related activity than alpha power alone^[26]^. In this study, the frontal alpha/beta ratio also showed a positive association with changes in positive affect after correction for multiple testing, suggesting that this measure may be a sensitive EEG correlate of the immediate positive affective response to CRM. In contrast, occipital IAF did not differ significantly among the three groups. This finding is informative because IAF reflects the peak frequency of the individual alpha rhythm and may be relatively stable over short time intervals. The lack of group difference in occipital IAF suggests that CRM did not substantially shift the individual alpha rhythm frequency in the immediate post-intervention period. Therefore, the short-term EEG response to CRM may be better characterized by changes in frontal arousal-relaxation balance and alpha-band functional connectivity than by changes in posterior alpha peak frequency^[27]^. The alpha-band wPLI findings further suggest that CRM may influence interregional functional coupling. The CRM group showed greater changes in alpha-band fronto-parietal and fronto-temporal wPLI than the control groups, indicating that CRM may modulate not only local frontal oscillatory activity but also coordinated activity between frontal and posterior or temporal regions. These networks may be involved in attention, bodily awareness, and affective processing. Although the connectivity findings should be interpreted cautiously because of the modest sample size, they support the possibility that the subjective relaxation response after CRM is accompanied by short-term network-level EEG modulation^[28]^.

### Multisensory mechanisms and the contextual role of cracking sound

The present findings help refine the interpretation of the immediate response to CRM by separating the cracking sound cue from the broader manipulation procedure^[29]^. In clinical practice, cracking sound is often interpreted as evidence that a joint release has occurred and that the manipulation has been successfully delivered. However, adding a simulated cracking sound to sham manipulation did not reproduce the full subjective or EEG response observed after real CRM. Although the sham + simulated sound condition retained a certain degree of treatment credibility, it did not generate CRM-like improvements in positive affect, comfort, satisfaction, or alpha-band functional connectivity. This result supports the interpretation that cracking sound may serve as a contextual or expectancy-related cue, but it does not appear sufficient to account for the complete short-term response induced by real CRM. This distinction is important because CRM is not merely an auditory event but a complex multisensory procedure^[30]^. Compared with sham manipulation, real CRM involves positioning the cervical segment near the restrictive barrier, applying traction, delivering a rapid small-amplitude thrust, and producing brief deformation of periarticular, muscular, fascial, and cutaneous tissues^[31]^. These mechanical events may activate cutaneous receptors, muscle spindles, joint mechanoreceptors, and deep cervical proprioceptive afferents^[32]^. The cervical region plays an important role in head–neck orientation, postural regulation, and multisensory integration with visual and vestibular information^[33]^. Therefore, the sensory consequences of CRM are unlikely to be limited to a local joint event; rather, they may provide a brief but salient afferent input capable of modifying bodily awareness, perceived tension, and arousal state^[34]^.

From this perspective, cracking sound and manipulation-related sensory input may act at different levels. Cracking sound is primarily an external auditory cue that may influence expectation, treatment credibility, and the participant’s judgment of whether a “real” manipulation has occurred. In contrast, the HVLA thrust provides direct tactile, proprioceptive, and deep somatosensory input from the cervical region^[32]^. These inputs may engage bottom-up sensory pathways and interact with top-down appraisal processes, potentially contributing to the subjective sense of release or relaxation frequently reported after CRM. The present study therefore does not exclude a psychological role of cracking sound; rather, it suggests that the auditory cue alone cannot substitute for the integrated sensory experience generated by real CRM.

Together with the EEG findings discussed above, this sensory-contextual interpretation suggests that the short-term relaxation response after CRM may arise from the interaction between manipulation-related afferent input and the participant’s cognitive appraisal of the treatment experience^[35]^. The observed frontal alpha/beta and alpha-band connectivity changes indicate that the subjective response was accompanied by measurable resting-state EEG modulation, but these neural effects were not reproduced by simulated cracking sound alone. Thus, the cracking sound should be understood as one component of the treatment context rather than as the principal mechanism underlying the immediate relaxation response.

These findings also have implications for clinical communication. If cracking sound is overemphasized as the key indicator of successful CRM, patients may mistakenly believe that the absence of a sound means treatment failure. A more appropriate explanation is that the therapeutic experience of CRM depends on the quality, safety, specificity, and sensory characteristics of the manipulation procedure as a whole. Clinicians may therefore need to frame cracking sound as a possible accompanying phenomenon rather than a necessary marker of effectiveness. Such communication may reduce unnecessary attention to the sound itself and help patients focus on comfort, safety, symptom response, and functional improvement.

For future mechanistic studies, this sensory-contextual framework provides a useful direction. Subsequent work could separate different components of the CRM experience more precisely, including auditory cues, tactile contact, passive cervical movement, end-range positioning, thrust-related proprioceptive input, vestibular contribution, and expectation^[36]^. Combining EEG with autonomic measures such as heart rate variability, skin conductance, and respiratory indices may further clarify whether the observed EEG changes reflect relaxation, reduced arousal, enhanced bodily awareness, or a combination of these processes.

## Limitations

This study has several limitations. First, the participants were healthy volunteers, so the findings should be interpreted as short-term neurophysiological and experiential responses rather than direct evidence of clinical efficacy. Second, EEG was recorded approximately 4 min after the intervention, and transient neural responses during or immediately after CRM may have been missed. Third, full participant blinding was not feasible because the HVLA thrust was perceptible, and expectancy effects cannot be completely excluded. Finally, the naturally occurring cracking sound in the CRM group was not experimentally controlled, and the modest sample size means that the equivalence-oriented analyses, brain–behavior correlations, and exploratory EEG findings require confirmation in larger clinical trials.

## Conclusion

In this single-session randomized controlled EEG study in healthy volunteers, CRM produced greater improvements in positive affect, comfort, and satisfaction than sham manipulation or sham manipulation plus simulated cracking sound. These subjective responses were accompanied by short-term EEG changes, particularly in frontal alpha/beta balance and alpha-band fronto-parietal and fronto-temporal functional connectivity. In contrast, simulated cracking sound alone did not reproduce the full subjective or EEG response observed after real CRM. These findings suggest that the immediate relaxation response after CRM may be more closely related to manipulation-associated sensory input than to the cracking sound cue alone.

## Supporting information

S1 File. CONSORT checklist

S2 File. the International Positive and Negative Affect Schedule-Short Form

S3 File. Improved version of the massage comfort scale

S4 File. EEG Key Parameters & QC Thresholds

S5 File. Preprocessing Pipeline

S6 File. Clinical Trial Protocol

S7 File. Data used to build graphs

## Acknowledgment

The authors would like to thank the medical staff of The Affiliated TCM Hospital of Guangzhou Medical University for their support and assistance to this research.

## Author contributions

**Conceptualization:** Yuanxun Lin.

**Data curation:** Meiling Yu.

**Formal analysis:** Yaoshuai Yu.

**Funding acquisition:** Junhua Li, Yikai Li.

**Investigation:** Yuanxun Lin, Fan Xue.

**Methodology:** Yuanxun Lin, Fan Xue, Lixin Tang.

**Project administration:** Jundan Wei.

**Resources:** Weixing Zhong.

**Software:** Yaoshuai Yu.

**Supervision:** Weixing Zhong.

**Validation:** Meiling Yu.

**Writing – original draft:** Yuanxun Lin, Weixing Zhong.

**Writing – review & editing:** Yaoshuai Yu.

## Data availability statement

All relevant data are within the manuscript and its Supporting Information files.

